# Hepatitis Delta Virus Reporting Requirements in the United States and Territories: A Systematic Review

**DOI:** 10.1101/2023.10.10.23296840

**Authors:** Milaveh Assadi-Rad, Brenda E. Acosta, Matthew C. Hesterman, Braden S. Fallon, Rachel L. Hill, Ethan W. Farnsworth, Bree Barbeau, Dede Vilven, Keisa M. Lynch, Melodie L. Weller

**Affiliations:** University of Utah, School of Dentistry, Salt Lake City, UT; Utah Department of Health and Human Services, Salt Lake City, UT; Salt Lake County Health Department, Salt Lake City, UT; University of Utah, Department of Gastroenterology and Hepatology, School of Medicine, Salt Lake City, UT; University of Utah, School of Medicine, Division of Pathology, Department of Microbiology and Immunology, Salt Lake City, UT

## Abstract

Hepatitis D virus (HDV) is a rare co-infection with hepatitis B virus. Currently, HDV is not a nationally notifiable disease in the United States. Only 55% of states and territories require HDV reporting, and most lack defined case definitions. Standardization of reporting requirements is crucial for monitoring HDV epidemiology.

## Introduction

Hepatitis D Virus (HDV) is classified as a rare infectious disease, affective less than 200,000 people in the United States (US) population^1^. However, accurate estimation of HDV prevalence is hindered due to limited testing and tracking. At the federal level, the Centers for Disease Control and Prevention (CDC) do not categorize HDV as a nationally notifiable disease^2^. Multiple studies have found that HDV testing is not regularly performed in people with hepatitis B virus (HBV) infections in the United States^3–5^. The lack of mandatory reporting to the CDC, limited testing in the HBV population, and inconsistent state-level reporting policies make it difficult to gauge HDV prevalence and track transmission changes. This study aimed to clarify the current HDV reporting requirements in the US.

The American Association for the Study of Liver Diseases (AASLD) has established guidelines for HDV testing^6^. Currently, the AASLD guidelines recommend HDV testing only in HBV patients with known risk factors for HDV, including those born in regions with high HDV prevalence, men who have sex with men (MSM), intravenous drug use (IVDU), patients with a history of HCV and HIV, multiple sexual partners, or elevated liver enzymes with limited detectable HBV DNA. The AASLD guidelines may lead to limited HDV testing in patients with HBV who do not present with or have unreported risk factors^7^. For those tested, AASLD recommends testing for anti-HDV antibodies and, subsequently, HDV RNA in patients who are HDV antibody positive (Fig 1)^6^.

**Figure 1.**
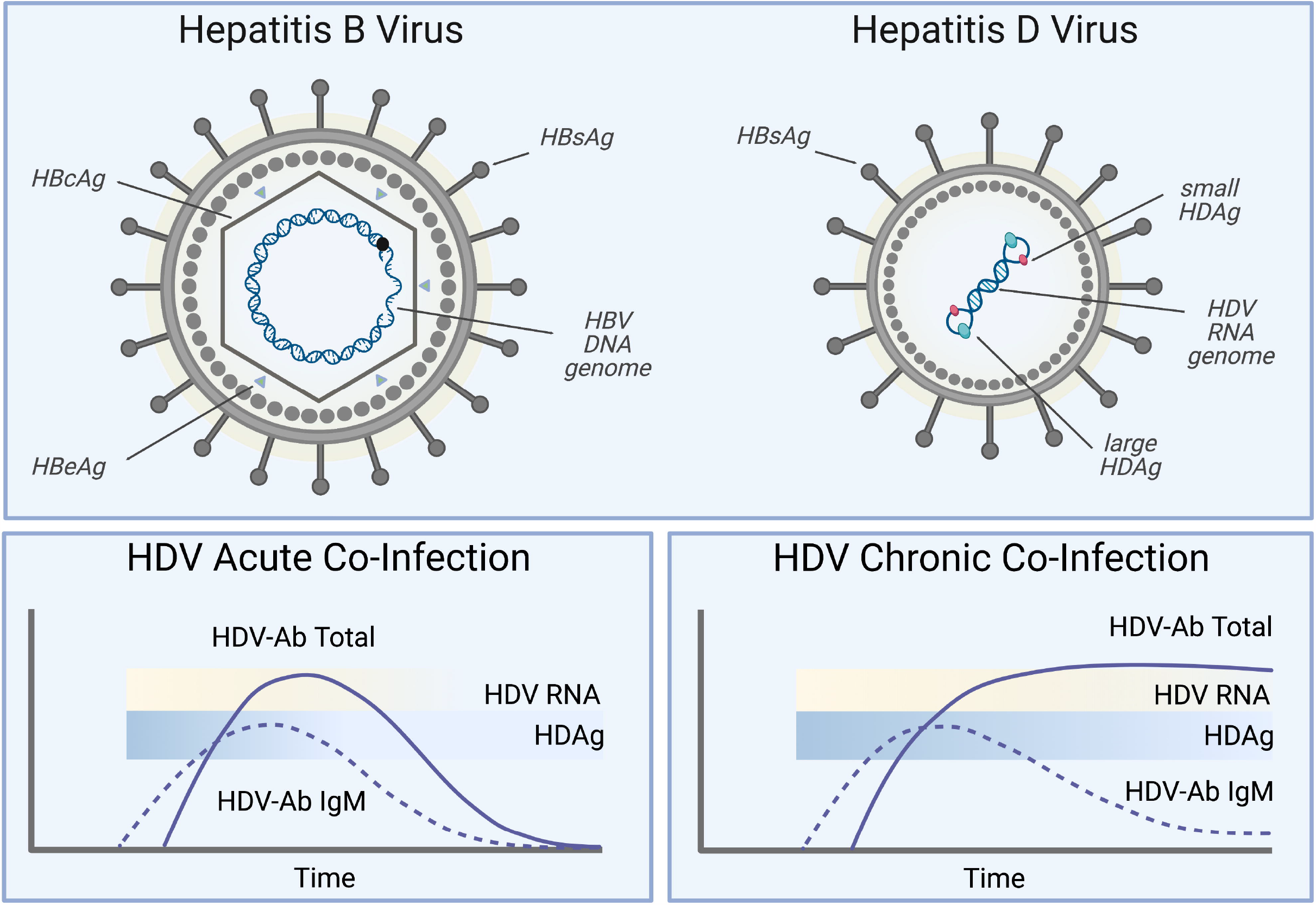
Hepatitis D Virus (HDV) and Hepatitis B Virus (HBV) structures and HDV testing guidelines. **A)** HDV is a satellite RNA that requires a helper virus for packaging and transmission. The HDV ribonucleoprotein complex, composed of the HDV RNA genome and two viral antigens (S-HDAg and L-HDAg), is packaged into an envelope membrane containing HBV surface antigens (HBSAg). HDV then utilize the same cell surface receptor as HBV, the sodium taurocholate co-transporting polypeptide (NTCP), and shares a similar tissue tropism profile as HBV. **B)** Testing for acute HDV coinfections is commonly performed with anti-HDAg IgM to detect recent HDV infections and anti-HDAg IgG or total anti-HDAg antibodies to detect acute HDV infections at later stages of infection. Patients who test positive for HDV-targeted antibodies should be tested for HDV-RNA. **C)** Testing for chronic HDV coinfections focuses on the detection of anti-HDAg IgG or total anti-HDAg antibodies. Similar to acute HDV co-infection testing, patients that test positive for HDV-targeted antibodies may further be tested for HDV-RNA and/or HDAg. Image was prepared in Biorender.

This study aimed to investigate and clarify the current requirements for HDV reporting in the United States and territories. The findings of this study revealed a lack of clear and consistent HDV reporting requirements and testing guidelines across the United States and territories. Current testing guidelines and limited tracking of HDV cases present a significant challenge in accurately estimating the prevalence of HDV and monitoring changes in HDV transmission patterns in the United States.

## Methods

### Evaluation of the United States and territory HDV reporting status

This study sought to determine the HDV reporting requirements for each state and territory in the United States. The state and territorial health departments were either queried through health department websites or contacted directly to obtain information on the HDV reporting requirements. The sources identified for states and territories that require reporting of HDV-positive lab results to public health departments are listed in Supplemental Table 1. The HDV reporting status may have been noted as Hepatitis D Virus, HDV, or other viral hepatitis to denote the required HDV reporting to the state or territorial health departments. The sources evaluated for the HDV reporting requirements in states that do not require HDV reporting are listed in Supplemental Table 2. Additionally, HDV reporting status for states and territories were also obtained or confirmed for a subset of states from the Council of the State and Territorial Epidemiologists (CSTE) State Reportable Conditions Assessment (SRCA) query tool (http://srca.querytool.cste.org/).

### Evaluation of required HDV testing for reporting diagnoses

In states that required HDV reporting, the specific laboratory tests required for reporting were queried. This information was obtained through state or territorial health department websites or departments were contacted directly to obtain information on HDV case definitions.

## Results

### 55% of United States and territories require reporting of HDV diagnoses

Of the 50 states, the District of Columbia, and the five territories of the United States, 55% (31/56) required reporting of HDV-positive lab results (Fig 2). Of the 50 United States and the District of Columbia, 29/51 (57%) required reporting of HDV cases to state health departments (Supplemental Table 1-2). Of the five United States territories (Guam, Northern Mariana Islands, Puerto Rico, American Samoa, and United States Virgin Islands), only Guam and Puerto Rico require the reporting of HDV cases. In addition to specific guidelines requiring the reporting of HDV cases, some states that do not specifically require HDV reporting may require reporting of “occurrence of any unusual disease of public health importance, “which may include HDV^8^. Together, 45% of the United States and territories currently do not require reporting of HDV cases.

**Figure 2.**
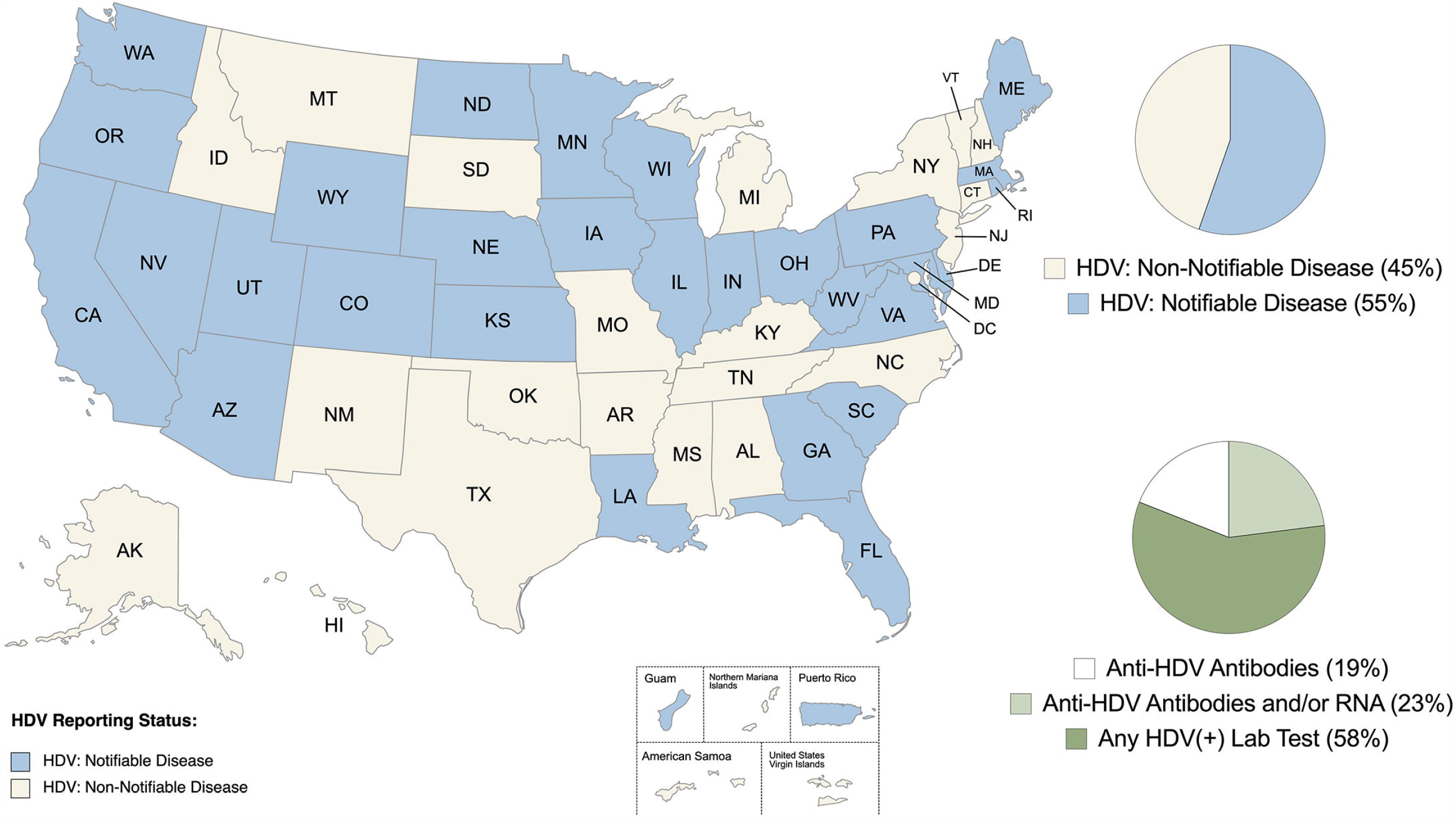
Map of Hepatitis Delta Virus Reporting Requirements for United States and territories. The map details states and territories that require reporting of HDV diagnoses. Thirty-one of the 56 United States and territories (55%) required the reporting of HDV-positive laboratory tests to the local health departments. Currently, 45% of the states and territories do not categorize HDV as a notifiable disease. Of the 31 states and territories requiring reporting of HDV, 19% required reporting of positive anti-HDV antibodies lab results, 23% required reporting of positive anti-HDV antibodies and/or HDV RNA lab results, and 58% reported accepting any HDV-positive lab results. Map produced in mapcharts.net

### HDV testing required for reporting HDV diagnoses

Of the 31 states and territories requiring reporting of HDV cases, readily accessible and detailed case definitions for HDV testing were not available for most of the states and territories. A majority of the states and territories (58%) would accept any HDV-positive laboratory results. Of the states and territories that had more defined HDV case definitions, 19% required reporting of positive anti-HDV antibody lab results and 23% required either anti-HDV antibody-and/or HDV RNA-positive lab results. The recommended tests for HDV detection may vary depending on the type of HDV infection being evaluated, including acute or chronic HDV co-infections (Fig 1). For the detection of HDV infections, the recommended tests are anti-HDV IgM, total anti-HDV, HDV-RNA or HDAg (transient)^6,9^.

## Discussion

This study aimed to evaluate the reporting requirements and testing protocols for HDV cases in the United States, District of Columbia, and five U.S. territories. The results indicated that 55% of the surveyed locations required HDV reporting. Among the states that required HDV testing, 58% lacked a clear HDV case definition and would accept any positive HDV lab result. This study underscores the need for clearer guidelines on HDV testing and reporting across the United States and territories. With only 55% of states and territories requiring the reporting of HDV-positive test results, it is crucial that HDV be added to the CDC’s list of notifiable infectious diseases to enable accurate tracking and characterization of HDV epidemiology nationwide.

Currently, limited information is available regarding the epidemiology of HDV in the United States, with only a few studies highlighting the insufficient testing of HDV in patient populations. Kushner et al. discovered that only 8.5% of HBV patients in the VA healthcare system were tested for HDV, with a positivity rate of 3.4%^3^. Wong et al. also found that 6.7% and 19.7% of HBV patients were tested for HDV in Quest Diagnostics and the VA healthcare system, respectively, and a positivity rate of 2.2% and 3.1% in the two independent cohorts tested, respectively^4^. Hesterman et al. found that 22% of HBV patients were tested for HDV, with a positivity rate of 8.3% in a Utahn patient cohort^5^. Other studies of HDV epidemiology in the United States have echoed the limited testing of HDV in the HBV patient population^7,10^. Fallon et al. evaluated international HDV epidemiology and reported significant structural breaks of unknown origin in HDV epidemiology^11^. These studies emphasize the need for increased surveillance of HDV.

HDV coinfections can have a significant impact on the progression of HBV-mediated disease, including higher risk of liver decompensation, hepatocellular carcinoma, and death, as well as a financial burden on the healthcare system^12,13^. The early detection of HDV in patients could enable early therapeutic intervention. The European Association for the Study of the Liver (EASL) and the Asian Pacific Association for the Study of the Liver (APASL) updated their HDV testing guidelines for HDV, HCV, and HIV testing in all HBV patients^14,15^. Palom et al. demonstrated a significant increase in HDV detection using updated EASL HDV testing guidelines^16^. Therefore, auto-reflex testing for HDV in all HBV patients and initiating standardized HDV case definitions and reporting requirements in the United States would support early detection of HDV and initiation of HDV therapeutics^17^.

The limitations of this study include updating of HDV reporting policies and limited accessibility to HDV case definitions. State-level policies on HDV reporting are frequently changing, and during manuscript preparation, some states and territories added or removed HDV reporting requirements. There may have been delays between reporting policy modifications and those reported via online reportable infectious disease websites for each state and territory.

The CSTE recommends the addition of diseases to the national notifiable disease list maintained by the CDC^18^. However, currently, there is no nationally defined HDV case definition and federal-level reporting requirements are absent. To better understand and track HDV in the United States, it is essential to establish clear and consistent guidelines for HDV testing and reporting at the state and territory levels. Ultimately, establishing a standardized HDV case definition and adding HDV to the CDC’s national notifiable disease list would provide a more comprehensive approach for monitoring HDV epidemiology in the United States and territories.

## Data Availability

All data produced are available online as noted in supplemental materials.

## Acknowledgements

On behalf of the authors, we would like to thank the University of Utah, School of Dentistry for their support throughout this study. We would also like to extend our gratitude to the state and territorial health departments for their cooperation and assistance in providing the data necessary for this study. We would like to thank the reviewers of this manuscript, including E.C. Diggins, T. Boone, and A. Romano. Finally, we acknowledge the editorial assistance provided by ChatGPT (OpenAI) for minor editing of the manuscript.

