## Supplemental Table 1 for "Hepatitis Delta Virus Reporting Requirements in the United States and Territories: A Systematic Review"

**Supplemental Table 1.** United States and territories requiring reporting of HDV cases.

| State or Territory | Health Department |
|---|---|
| Arizona (AZ) | Arizona Department of Health Services |
| California (CA) | California Department of PublicHealth |
| Colorado (CO)* | Colorado Department of Public Health and Environment |
| Delaware (DE)* | Delaware Division of Public Health |
| Florida (FL) | Florida Health |
| Georgia (GA) | GNR Public Health |
| Guam (GU) | Guam Department of Public Health and Social Services |
| Illinois (IL) | Illinois Department of Public Health |
| Indiana (IN) | Indiana State Department of Health |
| Iowa (IA) | IDPH Iowa Deparment of Public Health |
| Kansas (KS) | Kansas Department of Health and Environment |
| Louisiana (LA) | Louisiana Health department |
| Maine (ME) | Division of the Maine Department of Health and Human Services |
| Maryland (MD) | Maryland Department of Health |
| Massachusetts (MA) | Executive Office of Health and Human Services |
| Minnesota (MN) | Minnesota Department of Health |
| Nebraska (NE) | Nebraska Department of Health and Human Resources |
| Nevada (NV) | Nevada department of Health and Human Sevices |
| North Dakota (ND) | North Dakota Health Department |
| Ohio (OH) | Ohio Department of Health |
| Oregon (OR) | Oregon Health Authority |
| Pennsylvania (PA) | Pennsylvania Department of Health |
| Puerto Rico (PR)* | Puerto Rico Department of Health |
| Rhode Island (RI) | State of Rhode Island Department of Health |
| South Carolina (SC) | South Carolina Health Department |
| Utah (UT) | Utah Health Department |
| Virginia (VA)* | Virginia Department of Health |
| Washington (WA) | Washington State of Health Department |
| West Virginia (WV) | West Virginia Department of Health and Human Resources |
| Wisconsin (WI) | Wisconsin Department of Health Services |
| Wyoming (WY) | Wyoming Department of Health |

* Health Departments note required reporting of other viral hepatitis
