## Supplemental Table 2 for "Hepatitis Delta Virus Reporting Requirements in the United States and Territories: A Systematic Review"

**Supplemental Table 2.** United States and territories that do not require reporting of HDV cases.

| State or Territory | Health Department Source Hyperlinks: |
|---|---|
| Alabama (AL) | Alabama Public Health |
| Alaska (AK) | Alaska department of Health |
| American Samoa | American Samoa Department of Commerce |
| Arkansas (AR) | Arkansas Department Of Health |
| Connecticut (CT) | Connecticut State Department of Public Health |
| District of Columbia (DC) | District of Columbia Department of Health |
| Hawaii (HI) | State of Hawaii, Department of Health |
| Idaho (ID) | Idaho Department Health and Welfare |
| Kentucky (D) | Kentucky Cabinet for Health and Family Services |
| Michigan (MI) | Michigan Department of Health and Human Services |
| Mississippi (MS) | Mississippi State Department of Health |
| Missouri (MS) | Missouri Department of Health and Senior Services |
| Montana (MT) | Montana Department of Public Health and Human Services |
| New Hampshire (NH) | New Hampshire Department of Health and Human Services |
| New Jersey (NJ) | State of New Jersey Department of Health |
| New Mexico (NM) | New Mexico Department of Health |
| New York (NY) | New York Department of Health |
| North Carolina (NC) | North Carolina Department of Health and Human Services |
| Northern Mariana Island (MP) | CNMI Department of Public Health |
| Oklahoma (OK) | Oklahoma State Department of Health |
| South Dakota (SD) | South Dakota Department of Health |
| Tennessee (TN) | Tennessee Department of Health |
| Texas (TX) | Texas Health and Human Services |
| Vermont (VT) | Vermont Health Deparment |
| Virgin Islands (VI) | Virgin Islands Department of Health |
